# Evidence of Gender Disparities in Citations and Altmetric Attention Score in Oncology

**DOI:** 10.1101/2024.04.26.24306437

**Authors:** Rebecca A. Campbell, Emma Helstrom, Lauren Chew, Renu Eapen, Elizabeth Plimack, Andres Correa, Maarten Albersen, Alexander Kutikov, Philip Abbosh, Adam Calaway, Amanda Nizam, Shilpa Gupta, Sarah Psutka, Pedro Barata, Nazli Dizman, Mohit Sindhani, Christopher Weight, Georges Haber, Laura Bukavina

## Abstract

In this study, we assess gender disparities in both citation rates and Altmetric Attention Scores (AAS) within oncology, utilizing a dataset comprising over 600,000 articles from the past 15 years. The analysis focuses on the differential impact of gender on citation metrics and AAS, the latter of which expands the evaluation of research impact beyond traditional citation analysis to include digital engagement metrics such as social media mentions, news coverage, and policy discussions.

Methodologically, the study taps into the Altmetric database, using gender identification tools like the Gender Guesser API to classify articles by the gender composition of their first and last authors. Findings reveal a systemic over-citation of male-first and male-last author pairs (MM), with a mean citation difference (MCD) of +4.7. In stark contrast, female-first and female-last author pairs (WW) experience under-citation, with an MCD of -3.5.

A detailed examination of the Altmetric Attention Score shows that female-first authors receive a 7.19% lower AAS compared to male-first authors, indicating significant gender biases in the broader academic and public engagement. Delving deeper into the components of AAS, specifically news media coverage, reveals that female-first authors are covered 10.9% less in news media, 20.4% less patent references and 33.4% in Weibo mentions than their male counterparts. This disparity is significant and suggests a broader trend of underrepresentation and undervaluation of female-led research in more public-facing and influential media outlets.

These results underscore the persistent influence of gender on the recognition and valuation of scientific contributions within the field of oncology. They highlight the need for academia and publishing entities to implement more equitable practices to mitigate these disparities and ensure a balanced representation and recognition of scholarly work across genders.

## Introduction

Gender disparities in academia are a global concern, reflecting significant imbalances in the professional landscape of academic and research institutions.^1–3^ Among the various indicators of these disparities, the differences in citation counts— referred to as “citation disparity”—is particularly troubling.^4,5^ This phenomenon not only underscores potential biases in the recognition of scholarly contributions but also directly influences career advancement and the allocation of resources within academic communities.

Previous studies have indicated that publications authored by male researchers tend to garner more citations than those authored by females.^6–9^ This trend can be partly attributed to cognitive biases where male scholars are more likely to recall and cite works by their male peers. Further compounding the issue, research has shown that works with women listed as first or last authors receive approximately 30% fewer citations than comparable works authored by men in similar positions. This discrepancy manifests what is often termed the “Matthew Effect,” where male researchers’ contributions are seen as more central and pivotal to their fields. In contrast, the “Matilda Effect” describes the relative under-recognition and undervaluation of scientific achievements when women are the principal researchers.^10^

The persistent citation disparity between male and female researchers has prompted various explanations, including differences in resource allocation for open access publications, collaboration dynamics, academic rank, areas of specialization, and the topics addressed in published works.^11–13^ The continuation of these disparities may foster a simplistic view that the scholarly contributions of male and female scientists are fundamentally different. However, this perspective fails to consider the significant evidence of systemic barriers that impede women’s progress in academia. Previous research highlights the biases in perceptions of excellence, which can influence opportunities for visibility and advancement provided to individuals.^14^

While citations are traditionally seen as a key indicator of academic productivity and often serve as a basis for research and academic promotions, the introduction of the Altmetric Attention Score (AAS) provides a more nuanced measure of research impact. This score not only assesses the volume of citations but also evaluates the quality of attention that scholarly articles receive. It includes a range of complementary metrics, such as article downloads, mentions in policies and patents, media coverage, and engagement within professional networks (e.g., reference managers) and social media platforms like Twitter and mainstream media.^15^ This expansive approach offers a more comprehensive assessment of an article’s influence, presenting potential avenues to explore and address the intricate dynamics of citation and attention disparities in academia.^8^ Despite its potential, AAS remains underutilized in examining gender disparities.

In this study, we aim to analyze potential disparities in the Altmetric Attention Score specifically within the field of oncology over the past 15 years, spanning over 600,000 articles. We will examine the impact of research publications authored by female first and last authors compared to those authored by males. Furthermore, our study seeks to quantify the extent of over/under-citation in oncology literature, with an additional focus on differences across various subspecialties.

## Methodology

### Sample creation

The initial dataset was compiled from the Altmetric database through Application Programming Interface (API) obtained through Drexel University focusing on scholarly published articles that include specific keywords: ’sarcoma’, ’radiation therapy’, ’stem cell transplant’, ’targeted therapy’, ’immunotherapy’, ’melanoma’, ’malignancy’, ’lymphoma’, ’chemotherapy’, ’carcinoma’, ’neoplasm’, ’tumors’, ’oncology’, ’leukemia’, ’carcinogenesis’, ’metastasis’, ’adenocarcinoma’, ’squamous cell carcinoma’, ’glioma’, ’myeloma’, ’cancer genetics’, ’precision medicine’, ’cancer screening’, ’cancer prevention’, ’cancer diagnosis’, ’palliative care’, and ’cancer epidemiology’. The search was focused on articles published between January 1, 2009, and January 31, 2024. This approach yielded an extensive dataset comprising 652,834 records across the various oncology subfields.

In addition to the gender of the authors, our dataset was enriched with a variety of fields to support our analysis. These can be accessed in eMethods.

The dataset was rigorously preprocessed to maintain data accuracy and pertinence to our research objectives. This involved normalizing author names and correcting discrepancies within the metadata. To enhance the scope of papers suitable for gender classification based on first names, we utilized the DOI crossref API for comprehensive name citation matching.^16^ This process enabled us to associate abbreviated names, often represented by initials in certain journal entries, with their full-name counterparts. For instance, an entry such as L.B. Smith, accompanied by a DOI, was matched to its full form, Laura Bernice Smith, facilitating the application of the GenderGuesser API.^17^

### Name based assignment of author gender categories

In our dataset, we established “author gender categories” based on the first names of the first and last authors of the papers. Using the Gender Guesser API—a commercial tool providing statistical name-gender correlations across 191 countries— we assigned gender to authors whose first names had at least a 99.9% probability of being associated with a particular gender, labeling them as ’man’ or ’woman’ accordingly.

Our comprehensive dataset consisted of 652,843 papers, and we were able to confidently assign gender to both first and last authors in 75.1% of the papers. Furthermore, for 19.2% of the papers, gender could only be ascribed to either the first or last author. Papers were then categorized based on the gender labels of the first and last authors. Those with at first and last author labeled ’woman’ were categorized as WW, even if the other authors’ genders were undetermined. Conversely, if both first and last authors were labeled ’man’, the paper was categorized as MM. It is important to clarify that “author gender category” is a statistical tool and does not necessarily reflect the actual gender identities of the individuals. It is a proxy indicating a correlation between a given name and gender identity as commonly recognized on social media or official documents. The actual gender identity of authors could only be discerned through self-identification or personal knowledge, which lies beyond the scope of an automated analytical process such as ours.

Nonetheless, the use of author gender categories serves as a practical approximation for our study since names significantly influence the perception of gender identity. Such perceptions can affect judgments of scientific merit and, by extension, citation practices—regardless of the true gender identity of the authors in question. Furthermore, our study concentrated on the first and last authors due to a well-established norm in life sciences authorship. Typically, the first author of life science articles is the junior author responsible for conducting the research, whereas the last author is usually the senior author who conceptualizes and finances the research.

### AAS score

For the first part of our analysis, we focused on the Altmetric Attention Scores (AAS) for each publication, analyzing the scores in relation to the gender of the first and last authors. This evaluation was critical to determine if the gender of the authors influenced the amount of attention their research garnered across all altmetric variables. We employed an Ordinary Least Squares (OLS) regression approach to establish a model for investigating the dynamics between the logarithmically transformed AAS (independent variable) and the volume of Dimensions citations (dependent variable). In this construction, we accounted for variables such as the first author’s gender, open access (OA) status, and the publication year, enabling a measured examination of their collective influence on the citation count. Our regression framework incorporated an intercept to capture the baseline effect beyond the scope of the explanatory variables. A constant term was introduced, facilitating the interpretation of the model’s coefficients with respect to a null baseline of the independent variables. Upon fitting the OLS regression model, we evaluated the statistical significance of each coefficient to determine the impact of gender, OA status, and AAS on the predicted number of citations.

### Subspecialty Delineation

We compiled a roster of peer-reviewed publications across various oncological subspecialties for analysis, which includes sectors such as breast oncology, gastrointestinal oncology, gynecologic oncology, head and neck oncology, hematology-oncology, neuro-oncology, urological oncology, radiation oncology, general oncology, surgical oncology, and molecular oncology. These journals span the surgical, medical, and scientific domains within cancer research literature. The selection of journals was predicated on the availability of comprehensive data and their respective impact factors. To gauge citation impact, a baseline citation score of 1 was established, reflective of the significance of receiving at least one citation within the first two years post-publication. Each subspecialty was represented by a minimum of three journals. In instances where a journal was relevant to more than one category, it was retained only in the category where it was most contextually appropriate, thus avoiding redundancy. In total, 64 journals were classified into eleven categories, with 451,881 total articles from 2009 to 2024.

### Subspecialty Metrics

Calculation of over/under citation was calculated as a percent difference of observed citation from the gender-blind expectations model. Gender blind expectations using generalized additive model on the binomial outcomes [MM, WW] (GAM) previously described was utilized.^18^ Briefly, gender blind null model predicts the probability of citations based on publishing journal, categorization as a research, year of publication, combined number of authors in the dataset. For each article, one can define its over/under citation of each author gender category as the percent difference of observed citations from gender blind expectations. The degree to which papers authored by man-man (MM) pairs are over- or under-cited can be quantified using the formula (oMM −eMM)/eMM∗100 where oMM represents the actual observed number of citations received by MM papers from the citing articles, and eMM the predicted count of citations for MM papers, as forecasted by the gender-neutral model. Details regarding all these parameters and accompanying tests of significance can be found in documentation of the ‘mgcv’ package in R. ^19^

## Results

A total of 652,843 papers were retrieved using Altmetrics API. The distribution of our gender categories is as follows: ’UU’ (unknown-unknown) pairs were the most common at 162,751, followed by ’MM’ (man-man) pairs at 157,494, and ’WM’ (woman-man) pairs at 92,245. ’WW’ (woman-woman) pairs accounted for 61,329 papers, while ’MW’ (man-woman) pairs were present in 53,461 papers. Papers with one unknown gender paired with a known man (’UM’) were 42,678, and those paired with a known woman (’WU’) were 25,945. Lastly, ’MU’ (man-unknown) and ’UW’ (unknown-woman) pairs numbered 36,180 and 20,760 respectively.

### Gender differences in overall attention and source of attention

The analysis shows a notable difference in the overall Altmetric Attention Score (AAS) and its individual components between women and men serving in the first and last author positions (as shown in Fig 1G for first authors and Fig 1H for last authors). Women in the first author position have a 7.19% lower AAS compared to their male counterparts, a statistically significant difference (p<0.001). The discrepancy is slightly less for last authors, with women exhibiting a 4.9 % lower AAS than men (p=0.004).

**Figure 1.**
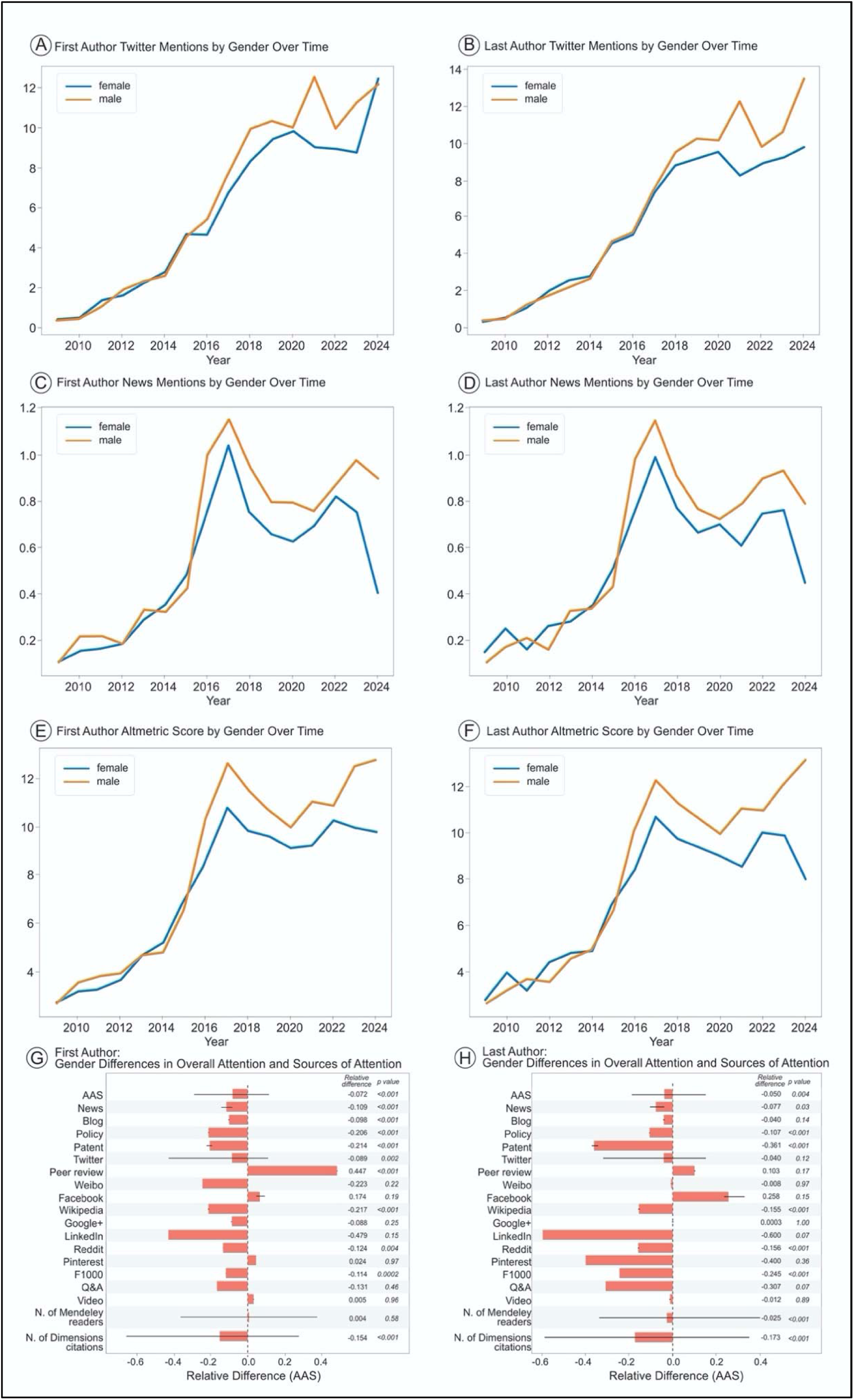
Analysis of Almetric Attention Scores (AAS) over time and by source of attention, including: A) first author Twitter mentions by gender over time; B) last author Twitter mentions by gender over time; C) first author news mentions by gender over time; D) last author news mentions by gender over time; E) first author AAS by gender over time; F) last author AAS by gender over time; G) relative differences in AAS for woman first author compared to male first author by specific source of attention; H) relative differences in AAS for woman last author compared to male last author by specific source of attention.

In terms of individual components measured by different sources of attention, first author women received less coverage by 10.9% in News (p<0.0001), 20.2% in policy discussions (p<0.0001), 20.4% in patent references (p<0.0001), 33.4% in Weibo mentions (p=0.05), and 8.2% in Twitter mentions (p=0.001). Similar trends were observed for women in the last author positions, with overall decreases in mentions across news (p=0.03), policy discussions (p<0.001), patents (p<0.001), Wikipedia references (p<0.001), and citation counts (p<0.001). Across all metrics within altmetrics, no single metric showed a statistically significant preferential mention of female first or last authors over males.

For the longitudinal analysis from 2009 to 2024, there has been an upward trajectory in engagement metrics across both gender lines. Nonetheless, our data reveal a persistent gender gap in the frequency of academic mentions. On social media, particularly Twitter, male authors outpaced female authors with an average of 12.5 mentions compared to 8.6 (p<0.001) (Fig 1A). A similar trend emerged in news media, with male authors receiving slightly more mentions than female authors by a margin of 0.2, peaking notably in 2022(Fig 1C). Further examination of altmetric score highlights a gender discrepancy favoring male first authors, who scored an average of 1.8 points above female first authors (Fig 1E).

Within last authorship, male last authors were more frequently mentioned in Twitter conversations, averaging 13.2 mentions in contrast to 9.5 for their female counterparts (p<0.001) (Fig 1B). This gender imbalance was echoed in news coverage, where male last authors were mentioned nearly twice as often as female last authors (0.8 vs 0.42 mentions, respectively; p<0.001) (Fig 1D). Additionally, the altmetric scores for 2024 reinforced this trend, with male last authors achieving scores averaging 12.5, significantly higher than the 8.1 average for female last authors (p=0.03) (Fig 1F).

### Citation Imbalance by Specialty

The proportion of papers in each author category (MM, WW, MW, WM) can be seen in Figure 2B as a function of publication. Note that percent of WM authorship represents the lowest total category with minimal growth from 2009 (10.2%) to 2024 (13.0%). Similarly, WW represent a small proportion of authorship, with minimal growth from 2009 (9.5%) to 2024 (14.4%)(Fig 2B).

**Figure 2.**
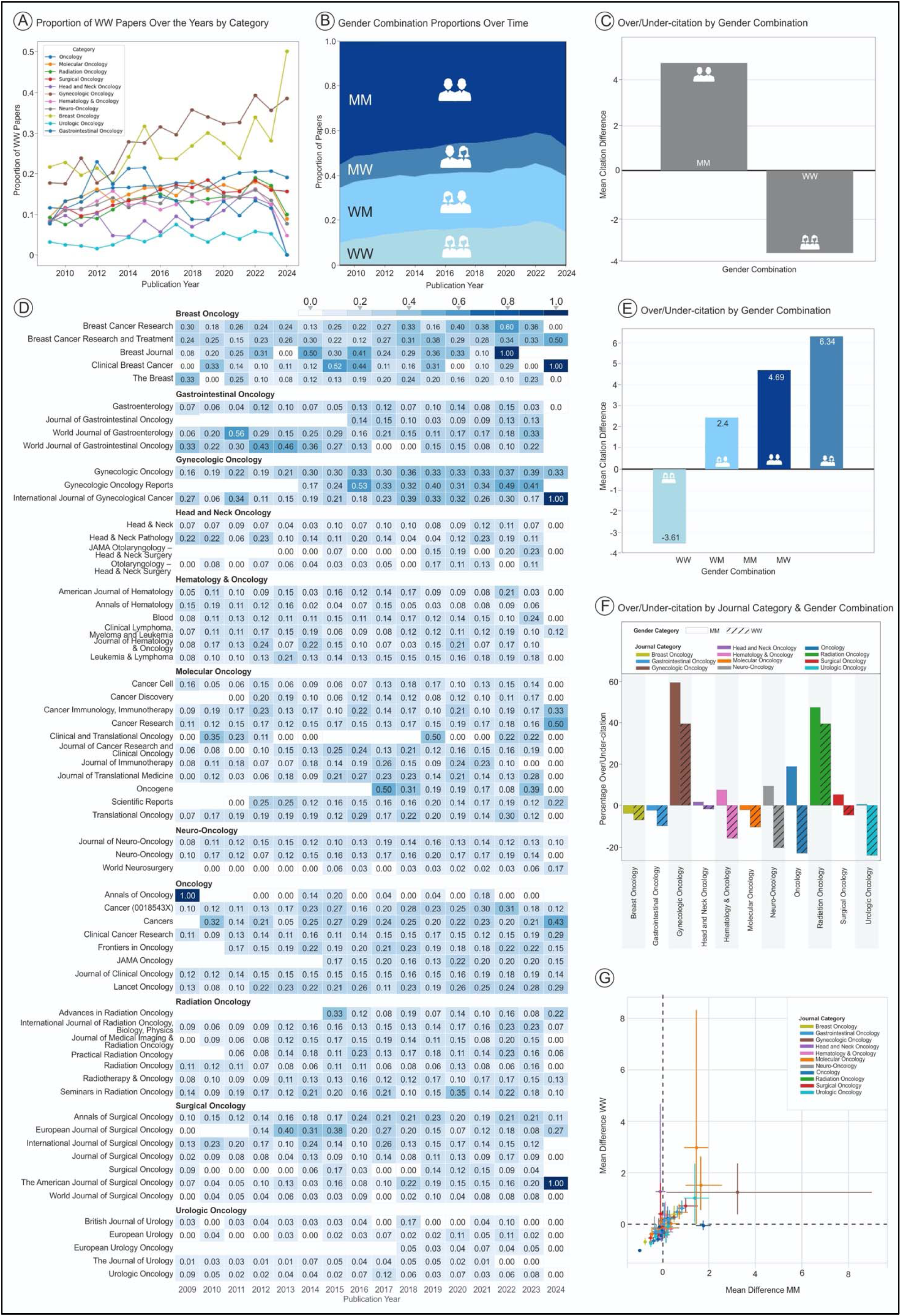
Analysis of gender combinations (gender of first author and gender of last author) and citations, including: A) proportion of WW papers over time by oncologic specialty; B) gender combination proportions over time (including WW, WM, MM, MW; C) over/under citation by gender combination (including MM and WW); D) proportion of WW papers over time by oncologic specialty (including specific high impact journals within each category); E) over/under-citation by all gender combinations (including WW, WM, MM, MW); F) over/under-citation by oncologic specialty and gender combination (including MM and WW); G) graph illustrating imbalance between mean difference in citations for WW versus MM with each point illustrating a specific journal which is color-coded according to the oncologic field in which it belongs. MM = male first author/male last author; MW = male first author/women last author; WM = women first author/male last author; WW = women first author/women last author.

The representation of WW authorship in oncology subspecialties has relatively modest growth, with the exception of Breast Oncology and Gynecologic Oncology, where there has been a marked increase in female authorship. Gynecologic Oncology, for instance, nearly doubled its representation of women authors, escalating from 17.7% in 2009 to 35.4% in 2023. Urologic Oncology, though starting with a lower baseline, saw its female authorship proportion rise from 3.1% to 5.2% in the same timeframe, reflecting a measurable and consistent growth. Overall, none of the subspecialties experienced a decline in women’s authorship, with the majority showing at least a nominal increase over the past fourteen years, as detailed in Figure 2A.

Figure 2D elucidates distinct inter-journal variations in the representation of women-first and women-last authorship across various oncology subspecialties. Notably, while the *Journal of Neuro-Oncology and Neuro-Oncology* maintain a comparable average of 13% for women authorship, *World Neurosurgery* exhibits a markedly lower figure, with women authors constituting less than 4.5% of its publications. In the realm of surgical oncology, the *Annals of Surgical Oncology* demonstrates a consistent inclusion of women in authorship roles, unlike the *World Journal of Surgical Oncology* and the journal *Surgical Oncology*, where women’s representation is significantly lower. These observations underscore the potential influence of individual journal policies and editorial practices on the prevalence of women’s authorship within their publications.

### Citation Imbalance

Our analysis of citation trends among gender combinations revealed strong evidence of gender bias. The MM combination has a striking over-citation value within oncologic academia, with a mean citation difference (MCD) of 4.7 (Figure 2C). In comparison, WW papers were found to be under-cited, with a MCD of -3.5. Furthermore, this trend generally persisted among all four gender combinations (Figure 2E). WW papers were the only cohort to experience under-citation (MCD -3.6). Interestingly, MW papers experience the highest over-citation with a MCD of 6.3 in comparison to MM with a MCD of 4.7. However, the overall pattern of WW papers being routinely under-cited and male authorship facilitating increased citation was observed. Dissecting these trends further, Figure 2F,G reveals that these citation gaps are not uniform across all subfields. In urologic oncology, MM experiences over-citation rate of 0.32%, while female-authored works are significantly undercited by 24.2%, bringing the total gender-based citation gap to 24.5%. Oncology stands out with the largest disparity, where MM pairs receive 19.0% more citations than anticipated, and WW pairs 22.9% fewer, cumulatively presenting a stark citation gap of 41.9%.

Nonetheless, the fields of gynecologic oncology and radiation oncology present an exception to this pattern. In these subspecialties, both WW and MM authorship configurations receive citations at rates exceeding expectations, with MM pairs at 59.5% and WW at 39.7% in gynecologic oncology, and 47.6% for MM and 39.5% for WW in radiation oncology. These exceptions indicate a potential for field-specific dynamics influencing citation practices, which may override the broader gender-based trends observed in citation behavior.

## Discussion

In our study encompassing an extensive dataset of 652,842 published manuscripts between 2009-2024, we sought to evaluate gender differences in proportions of papers published, citation metrics, and AAS across the field of oncology, an area which has not been extensively reported on in the literature. The data from our study reveal that publications authored by men, particularly those with male first and last authors (MM), are consistently cited more frequently than those authored by women. This trend was observed across various oncology subspecialties but was most pronounced in general oncology, where the gender citation gap was the largest. This could reflect a combination of cognitive biases where scholars are more likely to cite works from their male peers, as well as structural biases within the research community that might favor male principal investigators for clinical trials, research topics, or methodologies more commonly pursued by male researchers.

Interestingly, our results also showed that mixed-gender authorship pairs, specifically male first and female last authors (MW), experienced higher citation rates than expected. This suggests that the presence of a male author in a prominent position may mitigate some of the negative impacts of gender bias on citation rates. However, the fact that women-women (WW) pairs were consistently under-cited, even more so than mixed pairs, underscores the specific challenges faced by female scholars, especially when they occupy both pivotal roles in research authorship.

The under-citation of female-authored papers is troubling not only because it reflects bias but also because it directly impacts the visibility and influence of women’s research contributions. Lower citation rates have significant implications for career advancement, funding opportunities, and academic recognition. The “publish or perish” culture in academia places substantial weight on citation metrics. These metrics can influence a variety of professional outcomes, including graduate opportunities, funding success, career positions, awards, distinctions, and the critical processes of tenure and promotion.^20^ These trends have been substantiated by research, including a critical evaluation of the Altmetric Top 100 lists from 2015 to 2019, which demonstrated a significant citation gap disadvantaging female authors.^21^ Furthermore, an in-depth examination of more than 5000 articles from high-impact medical journals showed a recurring pattern of fewer citations for papers where women serve as first or last authors.^7^ Particularly pronounced was the finding that papers with women occupying both the first and last authorship positions experienced a citation rate halved compared to their male counterparts. These studies echo our own findings, pointing to an average gender citation gap of -8 in articles authored by women when contrasted with those authored by men, highlighting a critical issue that the academic community must address to promote equitable recognition of all researchers, and not just in theory but in practice.

Our study examined direct consequences of under citation utilizing the AAS, which has provided a detailed perspective on how research is perceived and valued across different platforms, including social media, policy documents, and news outlets.^8^ The AAS findings are indicative of a persistent gender gap when it comes to the visibility and recognition of academic work. Our results indicate that papers with women in the first author position were found to have an AAS that was 7.2% lower than those of their male counterparts—a significant difference that carries important implications for the reach and impact of their research. Last authors who are women also faced a deficit, with a 5.7% lower AAS compared to men.

More specifically, female first authors received lower coverage by 10.9% in news outlets, 20.2% in policy discussions, and 20.4% in patent references, with even more pronounced disparities observed in social media platforms such as Weibo and Twitter. Similar patterns were noted for women in last author positions, with reduced mentions and references across all examined media, suggesting a broad-based undervaluation of their contributions. In addition, over the fifteen-year period from 2009 to 2024, while we saw an overall increase in engagement metrics for academic work, the data underscore a gender divide, with male authors consistently receiving more mentions across the majority of platforms.

While there is a scarcity of research on AAS gender disparities specifically within oncology, other medical disciplines have begun to shed light on this phenomenon. Studies from cardiology, for instance, reveal that articles authored by women first authors are significantly less likely to achieve high AAS compared to those by men.^8^ Yet, when women occupy senior authorship roles, this gap in AAS does not seem to persist, and the difference in AAS between WW and men-men MM authored articles is not significant.

With the rise of social media as a pivotal platform for the dissemination of scientific and medical knowledge, the relevance of AAS and their associated sources of attention has surged, given their established correlation with citation rates. In the field of oncology, this correlation has been particularly noted in randomized phase III cancer trials, where a higher AAS is often an indicator of a greater number of citations.^22,23^ This trend is mirrored in gynecologic oncology research, where a robust positive relationship has been identified between median citation counts and median AAS.^24^ Moreover, evidence suggests a growing link between an article’s presence in high-impact journals and its AAS, implying that such publications tend to receive more media attention and, consequently, a higher citation rate. This correlation underscores the importance of amplifying the presence of female-authored research in prominent journals to bolster visibility and citation impact.^25^

This study faces several limitations. Primarily, the reliance on an online database to determine the gender of authors introduces the possibility of misclassification and incomplete data due to some authors being labeled as “unknown” or “excluded.” In addition, this approach does not account for the nuances of gender identity and self-identification, which transcend binary male or female categories. Such complexities are beyond the capacity of bioinformatical tools to discern, as they typically rely on name-based gender inference. The study also does not delve into the specifics of topics or publication types—such as videos or manuscripts—which can significantly influence AAS. For instance, studies have found that content related to treatment and quality of life, or the use of video abstracts, tends to attract more online engagement.^26^ Future research into the impact of gender on AAS might consider these factors more closely, including examining the gender makeup of citation sources and the proportion of female authorship in papers, which could provide deeper insights into the nuances of academic influence and recognition.

Despite existing challenges, there are several effective measures that can be implemented to narrow the persistent gaps in citation rates and AAS related to gender disparities. Establishing transparent and equitable citation practices is crucial. Journals can play a significant role by encouraging or requiring the inclusion of diverse work, particularly by integrating more women and underrepresented groups into their reviewer and editor pools to enhance visibility. Additionally, there should be a deliberate effort to promote the work of female researchers through social media and press releases, which could involve partnerships with academic institutions to showcase a broader range of research through various communication channels and support of open access publishing. Furthermore, funding bodies are encouraged to consider AAS and citation data within the context of existing biases when assessing grant applications. By developing policies that account for the influence of gender bias on these metrics, it’s possible to reduce their impact on funding decisions and support fairer research evaluation practices.

In conclusion, this manuscript has highlighted significant gender disparities in citation rates and AAS across various fields within oncology. Our findings highlight a systemic issue where female-authored research is under-cited and receives less attention compared to male-authored work, potentially impacting career advancement, funding opportunities, and academic recognition for women in science. As we advance, it will be essential to continue monitoring these metrics and implementing robust policies that not only recognize but actively combat the biases that perpetuate gender disparities in academic research.

## Data Availability

Data available upon request

## References

1. Mamtani M, Shofer F, Mudan A, et al. Quantifying gender disparity in physician authorship among commentary articles in three high-impact medical journals: an observational study. BMJ Open. Feb 25 2020;10(2):e034056. doi:10.1136/bmjopen-2019-034056

2. Fournier LE, Hopping GC, Zhu L, et al. Females Are Less Likely Invited Speakers to the International Stroke Conference: Time’s Up to Address Sex Disparity. Stroke. Feb 2020;51(2):674–678. doi:10.1161/STROKEAHA.119.027016

3. Boiko JR, Anderson AJM, Gordon RA. Representation of Women Among Academic Grand Rounds Speakers. JAMA Intern Med. May 01 2017;177(5):722–724. doi:10.1001/jamainternmed.2016.9646

4. Jagsi R, Guancial EA, Worobey CC, et al. The “gender gap” in authorship of academic medical literature--a 35-year perspective. N Engl J Med. Jul 2006;355(3):281–7. doi:10.1056/NEJMsa053910

5. Budrikis Z. Growing citation gender gap. Nature Reviews Physics. 2020/07/01 2020;2(7):346–346. doi:10.1038/s42254-020-0207-3

6. Filardo G, da Graca B, Sass DM, Pollock BD, Smith EB, Martinez MA. Trends and comparison of female first authorship in high impact medical journals: observational study (1994-2014). BMJ. Mar 02 2016;352:i847. doi:10.1136/bmj.i847

7. Chatterjee P, Werner RM. Gender Disparity in Citations in High-Impact Journal Articles. JAMA Netw Open. Jul 01 2021;4(7):e2114509. doi:10.1001/jamanetworkopen.2021.14509

8. Brown KN, Goel R, Soman S, et al. Gender Disparity in Citations and Altmetric Attention Scores in High-Impact Cardiology Journals. J Am Coll Cardiol. Aug 08 2023;82(6):572–573. doi:10.1016/j.jacc.2023.05.044

9. Ajay PS, Sharperson CM, Shah SK, Kooby DA, Shah MM. The Gender Gap in Surgical Literature: Are We Making Progress? J Surg Res. Mar 2024;295:357–363. doi:10.1016/j.jss.2023.11.033

10. Schmidt C. Matilda and Matthew Effects and Sexism in Science. In: Schmidt C, ed. The Invisible Hand of Cancer: The Complex Force of Socioeconomic Factors in Oncology Today. Springer International Publishing; 2023:121–133.

11. Lerchenmueller MJ, Sorenson O, Jena AB. Gender differences in how scientists present the importance of their research: observational study. BMJ. Dec 16 2019;367:l6573. doi:10.1136/bmj.l6573

12. Lerchenmüller C, Lerchenmueller MJ, Sorenson O. Long-Term Analysis of Sex Differences in Prestigious Authorships in Cardiovascular Research Supported by the National Institutes of Health. Circulation. Feb 20 2018;137(8):880–882. doi:10.1161/CIRCULATIONAHA.117.032325

13. Bendels MHK, Müller R, Brueggmann D, Groneberg DA. Gender disparities in high-quality research revealed by Nature Index journals. PLoS One. 2018;13(1):e0189136. doi:10.1371/journal.pone.0189136

14. Llorens A, Tzovara A, Bellier L, et al. Gender bias in academia: A lifetime problem that needs solutions. Neuron. Jul 07 2021;109(13):2047–2074. doi:10.1016/j.neuron.2021.06.002

15. Lerchenmueller MJ, Schmallenbach L, Bley M, Lerchenmüller C. Gender disparities in altmetric attention scores for cardiovascular research. Communications Biology. 2023/07/17 2023;6(1):741. doi:10.1038/s42003-023-05058-9

16. API C. https://github.com/CrossRef/rest-api-doc

17. API GG. https://gender-guesser.com/

18. Teich EG, Kim JZ, Lynn CW, et al. Citation inequity and gendered citation practices in contemporary physics. Nature Physics. 2022/10/01 2022;18(10):1161–1170. doi:10.1038/s41567-022-01770-1

19. Wood SN. Fast stable restricted maximum likelihood and marginal likelihood estimation of semiparametric generalized linear models. Journal of the Royal Statistical Society: Series B (Statistical Methodology). 2011;73(1):3–36. doi: 10.1111/j.1467-9868.2010.00749.x

20. Davies SW, Putnam HM, Ainsworth T, et al. Promoting inclusive metrics of success and impact to dismantle a discriminatory reward system in science. PLoS Biol. Jun 2021;19(6):e3001282. doi:10.1371/journal.pbio.3001282

21. Rotenstein LS, Torre M, Cleary JL, Sen S, Guille C, Mata DA. Differences in Gender Representation in the Altmetric Top 100. J Gen Intern Med. Feb 2022;37(3):590–592. doi:10.1007/s11606-021-06829-y

22. Rooney MK, Sharifi B, Ludmir EB, Fuller CD, Warner JL. Factors Associated With Altmetric Attention Scores for Randomized Phase III Cancer Clinical Trials. JCO Clin Cancer Inform. Aug 2023;7:e2300082. doi:10.1200/CCI.23.00082

23. Abi Jaoude J, Kouzy R, Rooney M, et al. Impact factor and citation metrics in phase III cancer trials. Oncotarget. Aug 31 2021;12(18):1780–1786. doi:10.18632/oncotarget.28044

24. Chi AJ, Lopes AJ, Rong LQ, Charlson ME, Alvarez RD, Boerner T. Examining the correlation between Altmetric Attention Score and citation count in the gynecologic oncology literature: Does it have an impact? Gynecol Oncol Rep. Aug 2021;37:100778. doi:10.1016/j.gore.2021.100778

25. Huang W, Wang P, Wu Q. A correlation comparison between Altmetric Attention Scores and citations for six PLOS journals. PLoS One. 2018;13(4):e0194962. doi:10.1371/journal.pone.0194962

26. Bonnevie T, Repel A, Gravier FE, et al. Video abstracts are associated with an increase in research reports citations, views and social attention: a cross-sectional study. Scientometrics. 2023;128(5):3001–3015. doi:10.1007/s11192-023-04675-9

